# No genetic evidence for *SLC7A11* involvement in Parkinson’s Disease

**DOI:** 10.64898/2026.07.01.26357025

**Authors:** Yoomin Lee, Sitki Cem Parlar, Emma Somerville, Konstantin Senkevich, Meron Teferra, Ziv Gan-Or

## Abstract

**Background:** The cystine/glutamate antiporter encoded by *SLC7A11* maintains cellular redox balances and lysosomal pH. Recent molecular evidence suggested that *SLC7A11* may be involved in Parkinson’s disease (PD). However, the genetic contribution of *SLC7A11* to PD susceptibility remains unclear.

**Methods:** We analysed whole-genome sequencing data from the Accelerating Medicines Partnership-Parkinson’s Disease (AMP-PD) and United Kingdom Biobank (UKBB) cohorts, comprising of 5,5375 cases and 35,002 controls. Rare variant burden analyses were performed using SKAT-O and MetaSKAT. Common-variant associations with PD risk and glucocerebrosidase (GCase) enzyme activity were assessed with regional linkage disequilibrium plots using previous GWAS summary statistics. Gene expression effects were examined through brain-specific expression quantitative trait loci (eQTL) data from GTEx v7. Colocalization analyses and regional association plots were generated to visualize and compare GWAS and eQTL signals across the SLC7A11 locus.

**Results:** No rare or common *SLC7A11* variants were associated with PD or GCase activity, including variants that had strong effects on SLC7A11 expression in relevant brain regions.

**Discussion:** These findings suggest that genetic variation in *SLC7A11* or its expression are unlikely to have a major role in PD susceptibility.

**Plain Language Title:** We did not find evidence that the *SLC7A11* gene is involved in Parkinson’s disease

**Plain Language Summary:** Parkinson’s Disease (PD) is a brain disorder where both genetic and environmental factors play a role in a person’s risk of developing PD. Recent studies suggested that a gene called *SLC7A11* may be involved in PD as it helps to decrease the factors that increase the chances of getting PD, such as cell stress and lysosome function.

In this paper, we tested the genetic importance of *SLC7A11* and PD by analysing data from two data sets: the Accelerating Medicines Partnership-Parkinson’s Disease study and the UK Biobank. Together, we studied both rare and common genetic changes of SLC7A11 in approximately 5,300 people with PD and 35,000 without. We also looked at whether these genetic changes affected GCase, an enzyme important for lysosomal function, and whether there was a difference in *SLC7A11* activity in different brain regions.

We found no evidence that genetic changes in *SLC7A11* were associated with PD risk or GCase enzyme activity. Some genetic changes affected *SLC7A11* expression in the brain, but these changes were not linked with PD.

Overall, our findings suggest that inherited genetic differences in *SLC7A11* are unlikely to play a major role in PD risk.

## 1. Introduction

Parkinson’s disease (PD) is a complex neurodegenerative disorder influenced by both genetic and environmental factors. Genome-wide association studies (GWAS) have identified numerous loci linked to PD risk, highlighting the importance of genetic susceptibility in disease development ^1^. Among the cellular pathways implicated by these genetic findings, lysosomal dysfunction has emerged as a central mechanism in PD pathogenesis ^2^. For example, variants in the lysosomal potassium/proton channel *TMEM175* have been associated with PD, with evidence suggesting that these variants may affect pH in the lysosome and reduce GCase activity ^3,4^.

*SLC7A11* encodes a cystine/Glu permease and transporter in lysosomal membranes which, like *TMEM175*, regulates lysosomal pH. *SLC7A11* has recently been suggested to be involved in PD, as its dysfunction seems to increased oxidative stress and over-acidification of the lysosome, resulting in an accumulation of alpha-synuclein ^5^. However, the potential genetic association between *SLC7A11* and PD at the population-level remains unclear.

In this study, we examined whether common and rare variants in *SLC7A11* are associated with PD risk, and if they affect RNA expression and glucocerebrosidase (GCase) activity.

## 2. Methods

For this analysis, we collected whole genome sequencing data from two datasets: the Accelerating Medicines Partnership in Parkinson’s Disease (AMP-PD) and the United Kingdom Biobank (UKBB). We then performed dataset-specific quality control procedures. For the UKBB data, the BCFtools package was used to exclude multi-allelic variants ^6^. We then filtered the data to retain only the high-quality data using the Genome Analysis Toolkit (GATK) v4.2.5 (https://github.com/broadinstitute/gatk.git) with parameters including genotype quality (GQ) ≥ 25 and read depth (DP) ≥ 25, with an exclusion of variants with > 5% missingness. For the AMP-PD dataset, no further filtering was done as the data came pre-processed and quality controlled according to the procedures detailed on their website (https://amp-pd.org/whole-genome-data). Finally, variants were filtered to retain only rare alleles, defined as those with a minor allele frequency (MAF) < 0.01.

For the UKBB cohort, PD cases were identified through four criteria: self-report, ICD-10 diagnosis codes, primary and secondary cases of death records, and algorithmically defined cases. We excluded atypical parkinsonism causes, including multiple system atrophy and progressive supranuclear palsy. Controls were selected after excluding all PD cases, proxy cases (defined as first degree relatives with a family history of PD), and individuals with movement disorders or neurodegenerative conditions. We also applied sample-level quality control filters, retaining only unrelated individuals (no closer than third-degree relatives), individuals of European ancestry, excluding those with sex chromosome aneuploidy or heterozygosity/missing outliers. Age- and sex-matched controls were then selected at a 10:1 control-to-case ratio using a 5-year binning method.

Following the application of quality control, analyses were conducted across both datasets, amassing a combined cohort of 5,375 cases and 35,002 controls (Table 1). Variants within the SLC7A11 locus were grouped into 5 functional categories, including all rare variants, non-synonymous variants, the top 1% of predicted deleterious variants (CADD ≥ 20), AlphaMissense predicted likely pathogenic, and loss-of function variants (stopgain, frameshift, and splice-site variants). Burden testing of the five *SLC7A11* variant categories was performed using optimized sequence kernel association tests (SKAT-O), followed by a meta-analysis across cohorts using MetaSKAT to examine the association of rare *SLC7A11* variants with PD. For the UKBB cohort, models were adjusted for sex and age, and the first 10 principal components to account for population stratification. For the AMP-PD cohort, models were adjusted for sex and age, as principal components were not available. Individual variant-level association was assessed using Fisher’s exact test, with odds ratios and 95% confidence intervals calculated for each variant. To account for multiple testing, false discovery rate (FDR) correction was applied using the Benjamini-Hochberg method.

**Table 1.** Demographic characteristics of the study cohorts.

| Cohort | Cases (N) | Case age,<br>mean (SD) | Male cases, n<br>(%) | Controls<br>(N) | Control age,<br>mean (SD) | Male controls,<br>n (%) |
| --- | --- | --- | --- | --- | --- | --- |
| UKBB | 3,444 | 70.35 (8.91) | 2,200 (63.9) | 31,940 | 72.20 (6.82) | 20,500 (64.2) |
| AMP-PD | 1,931 | 64.56 (9.49) | 1,245 (64.5) | 3,062 | 69.79 (13.03) | 1,486 (48.5) |
| All cohorts | 5,375 | 68.27 (9.54) | 3,445 (64.1) | 35,002 | 71.99 (7.60) | 21,986 (62.9) |

**Abbreviations:** AMP-PD, Accelerating Medicines Partnership – Parkinson’s Disease; n (%), number and percentage; N, number of individuals; SD, standard deviation; UKBB, UK Biobank

To examine the association of common variants with PD, we generated regional Manhattan plots using LocusZoom to visualise GWAS associations with linkage disequilibrium (LD) information ^7^. Analyses were specifically focused on the *SLC7A11* locus to assess common variant signals within the region. Similar methods were used study the association of this gene with glucocerebrosidase (GCase) activity. We obtained GWAS summary statistics for GCase from the GWAS Catalog ^8^. The genomic coordinates of gene *SLC7A11*, extended by 500,000 base pairs both upstream and downstream, were examined for significant association signals in this dataset following multiple testing correction. The data was then visualised using a regional association plot using LocusZoom.

In order to assess the effects of common variants in the *SLC7A11* locus on gene expression, we analysed the expression quantitative trait loci data (eQTL) from the Genotype-Tissue Expression (GTEx) Project, Analysis Release v7 ^9^. All available brain-related tissues were used in this study. We applied Bonferroni correction for multiple comparisons.

We performed a colocalization analysis using the coloc R package to identify any shared genetic signals between PD GWAS and *SLC7A11* eQTLs from GTEx v7 across various brain tissues ^7,10^. Summary statistics were harmonized using the LiftOver tool to convert GWAS coordinated from GRCh38 to GRCh37, aligning them with the GTEx v7 reference build ^11^. We then restricted the variants to a ±500 kb window around the *SLC7A11* locus. Analyses were conducted under an approximate Bayes factor framework with the GWAS modeled as a case-control trait and eQTLs as quantitative traits. Posterior probabilities were computed for five hypotheses (H0-H4), with colocalization defined as posterior probability of H4 (PP.H4) ≥ 0.8.

To complement the statistical colocalization, regional association plots were generated using the LocusCompareR package to visualize the genomic context of GWAS signals at the SLC7A11 locus ^12^. Variants within the ±500 kb region of *SLC7A11* were plotted according to their genomic position and negative log-transformed p-values to show the spatial relationship between PD-associated variants and eQTL signals across tissues.

## 3. Results

We first examined the association of rare variants in *SLC7A11* in PD using SKAT-O in each individual cohort, followed by a meta-analysis using metaSKAT. In the SLC7A11 locus, we identified 2,107 rare variants in the UKBB cohort and 602 in the AMP-PD cohort. The majority of variants were non-coding and no loss-of-function variants were identified in both datasets (Supplementary Data 1 & 2). No associations were found in both AMP-PD and UKBB cohorts (Table 2). We then examined the association of common variants in and around *SLC7A11* with PD using a regional association plot derived from the recent PD GWAS, and no statistically significant variants were found to be associated with PD risk (Figure 1, ^7^).

**Figure 1.**
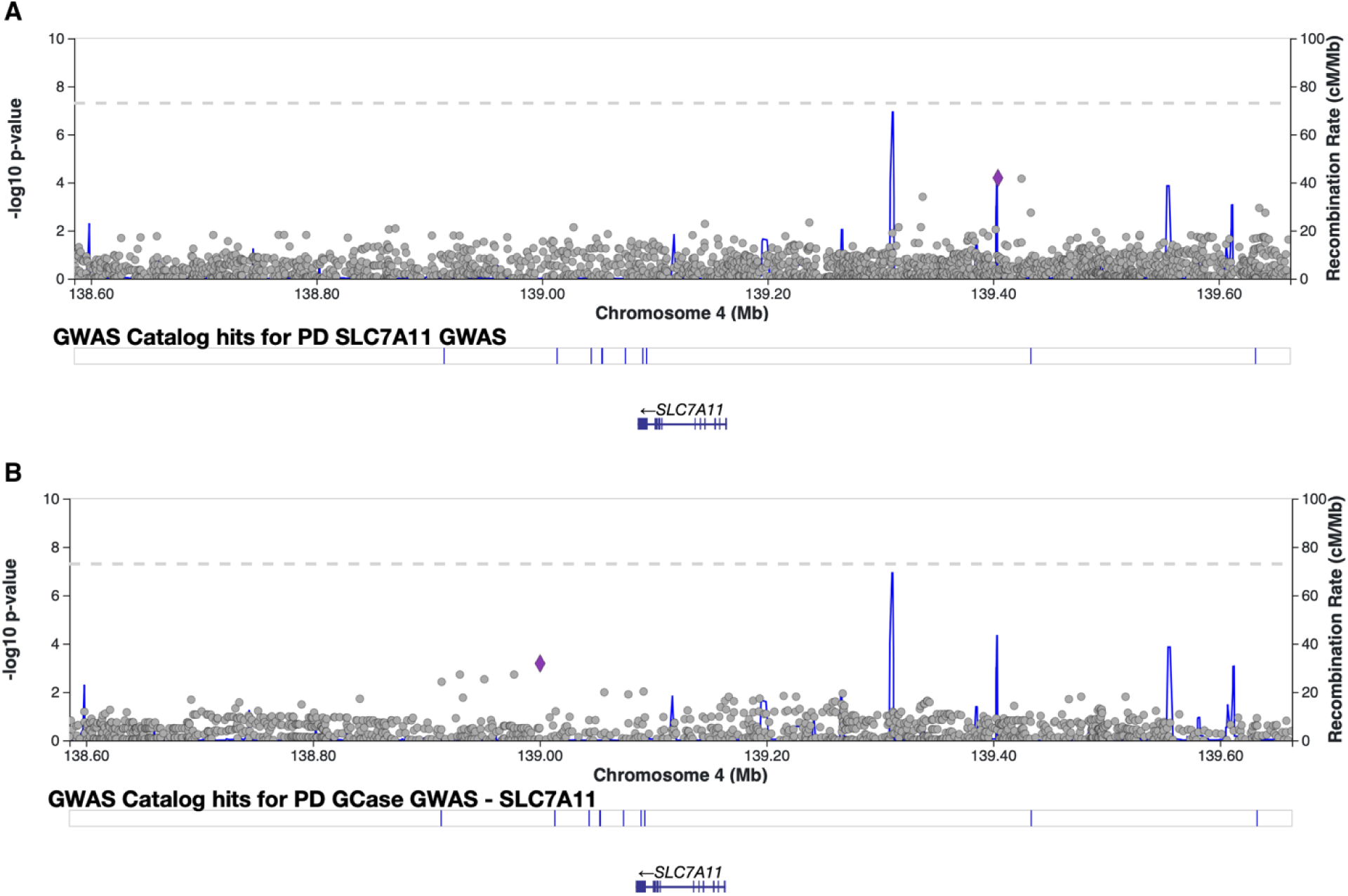
Regional association plots of common variants at the *SLC7A11* locus. (A) Regional association plot of common variants within the *SLC7A11* locus and Parkinson’s disease risk from the latest PD GWAS. (B) Regional association plot of common variants within the *SLC7A11* locus and glucocerebrosidase (GCase) enzymatic activity. The dashed grey line indicates the genome-wide significance threshold (p = 5 × 10-8). The purple diamond indicates the lead variant. Recombination rates (cM/Mb) are plotted on the right y-axis, indicated by the blue lines.

**Table 2.** Rare variant SKAT-O burden results for SLC7A11 across AMP-PD, UK Biobank, and MetaSKAT datasets in relation to Parkinson’s disease.

| Dataset | Variant set | p-value | FDR | N SNPs |
| --- | --- | --- | --- | --- |
| AMP-PD | *All rare* | 0.5577 | 1.0000 | 361 |
| AMP-PD | *CADD* | 0.4651 | 1.0000 | 10 |
| AMP-PD | *Non-synonymous* | 0.7682 | 1.0000 | 13 |
| AMP-PD | *AM* | 1.0000 | 1.0000 | 3 |
| UKBB | *All rare* | 0.5950 | 1.0000 | 533 |
| UKBB | *CADD* | 0.5409 | 1.0000 | 2 |
| UKBB | *Non-synonymous* | 0.5409 | 1.0000 | 2 |
| UKBB | *AM* | NA | NA | NA |
| MetaSKAT | *All rare* | 1.0000 | 1.0000 | – |
| MetaSKAT | *AM* | 1.0000 | 1.0000 | – |
| **MetaSKAT** | *CADD* | 1.0000 | 1.0000 | – |
| **MetaSKAT** | *Non-synonymous* | 1.0000 | 1.0000 | – |

**Abbreviations:** AM, AlphaMissense predicted likely pathogenic; AMP-PD, Accelerating Medicines Partnership – Parkinson’s Disease; CADD, combined annotation dependent depletion; FDR, false discovery rate; n (%), number and percentage; N, number of individuals; MetaSKAT, meta-analysis for SNP-Kernel Association Test; SD, standard deviation; SNP, single nucleotide polymorphism; UKBB, UK Biobank

We assessed common SLC7A11 variants for associations with GCase activity to investigate if this locus influences lysosomal enzyme function (Figure 2). No associations were significant after applying Bonferroni correction (p < 2.3 × 10 ^4^).

**Figure 2.**
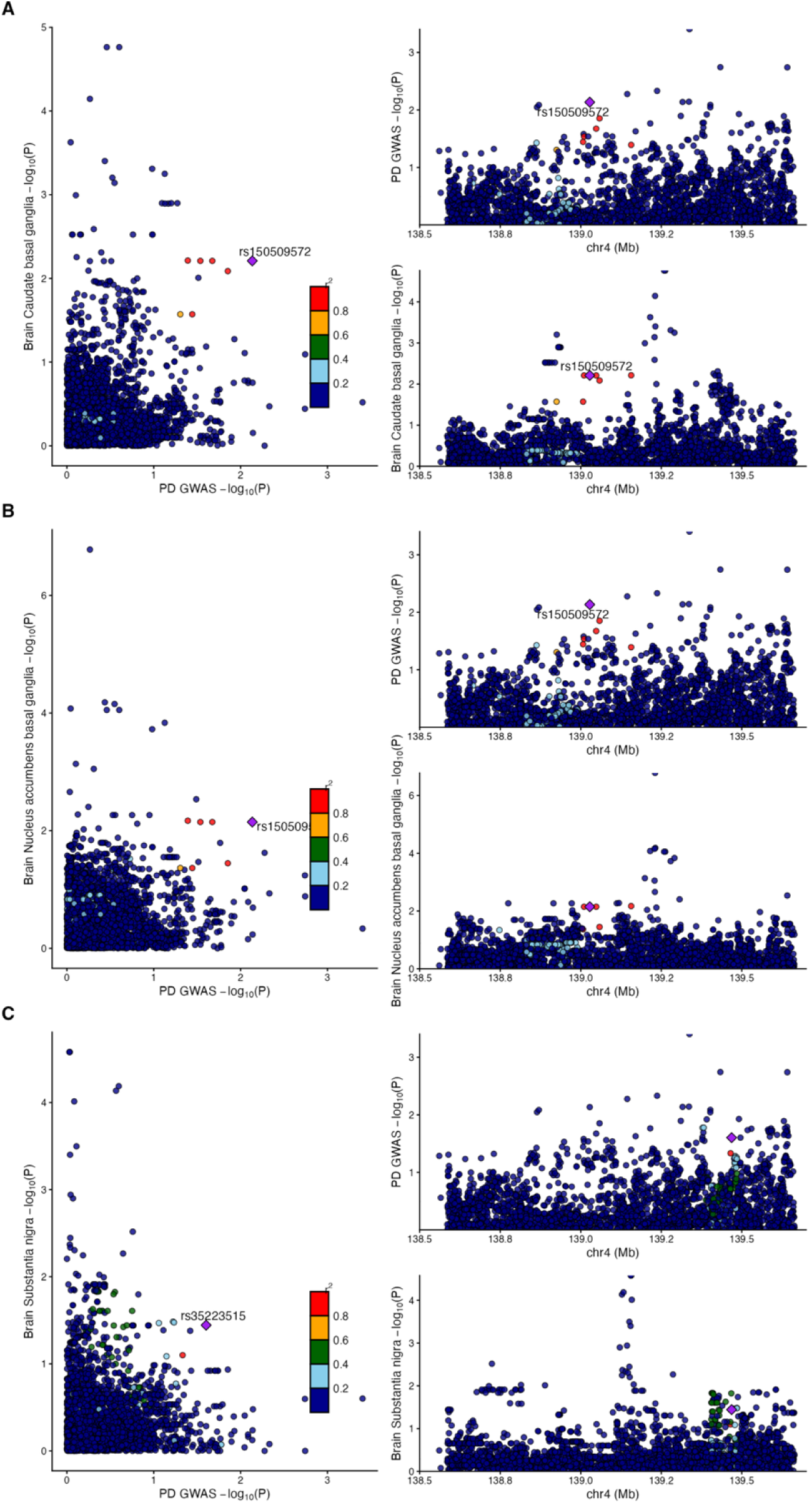
Colocalization of PD GWAS and *SLC7A11* eQTL signals in basal ganglia and substantia nigra. Scatterplots (left) and reginal association plots (right) comparing PD GWAS and *SLC7A11* eQTL -log_10_(P) values for variants within ±500 kb of the SLC7A11 locus, (A) Caudate (basal ganglia). (B) Nucleus accumbens (basal ganglia). (C) Substantia nigra. In the scatter plots, each point represents a shared variant, colored by linkage disequilibrium (r^2^) with the lead variant indicated as a purple diamond.

Next, we analysed eQTLs to determine whether variants within the *SLC7A11* locus were associated with gene expression in brain tissues. Eleven variants across multiple brain tissues were significant after multiple-testing correction (Bonferroni threshold p < 1.2 × 10, Supplementary Data 3). Notably, variant rs4863780 demonstrated the strongest association compared to the other variants and was consistently detected across several brain regions (Table 3). In the PD GWAS, this variant was not significantly associated with disease risk (p = 0.5426).

**Table 3.** SLC7A11 eQTLs with the strongest associations in brain tissues from GTEx.

| Brain Tissue | Variant | $\beta$ | SE | p-value |
| --- | --- | --- | --- | --- |
| **Nucleus accumbens (basal ganglia)** | rs4863780 | 1.40 | 0.09 | $5.49 \times 10^{-30}$ |
| **Caudate (basal ganglia)** | rs4863780 | 1.25 | 0.09 | $7.14 \times 10^{-29}$ |
| **Cortex** | rs4863780 | 0.95 | 0.07 | $3.54 \times 10^{-24}$ |
| **Putamen (basal ganglia)** | rs4863780 | 1.31 | 0.10 | $1.34 \times 10^{-23}$ |
| **Frontal Cortex (BA9)** | rs4863780 | 1.04 | 0.08 | $1.64 \times 10^{-23}$ |
| **Cerebellum** | rs4863780 | 1.27 | 0.11 | $1.69 \times 10^{-21}$ |
| **Hypothalamus** | rs4863780 | 1.27 | 0.10 | $1.09 \times 10^{-20}$ |
| **Cerebellar Hemisphere** | rs4863780 | 1.20 | 0.11 | $4.03 \times 10^{-19}$ |
| **Nucleus accumbens (basal ganglia)** | rs73850513 | 1.17 | 0.11 | $5.21 \times 10^{-19}$ |

**Abbreviations:** SE, standard error

Finally, to investigate whether PD association at the *SLC7A11* locus is mediated by changes in gene expression, we performed a colocalization analysis across twelve brain tissues. There was no evidence of a shared genetic signal between PD risk and *SLC7A11* expression in any of the brain tissues examined (Table 4). Across all analyzed tissues, the posterior probability of a shared causal variant (PP.H4) remained consistently low, at about 0.005, well below the predefined significance threshold of 0.80.

**Table 4.** Colocalization analysis of *SLC7A11* eQTLs and Parkinson’s disease GWAS across brain tissues.

| Brain Tissue | Number of SNP | H0 | H1 | H2 | H3 | H4 |
| --- | --- | --- | --- | --- | --- | --- |
| Caudate (basal ganglia) | 3850 | 0.6668 | 0.0504 | 0.2582 | 0.0195 | 0.0051 |
| Nucleus accumbens (basal ganglia) | 3849 | 0.6589 | 0.0496 | 0.2663 | 0.0201 | 0.0051 |
| Anterior cingulate cortex (BA24) | 3839 | 0.6451 | 0.0483 | 0.2807 | 0.021 | 0.005 |
| Cerebellar hemisphere | 3852 | 0.6732 | 0.051 | 0.2518 | 0.0191 | 0.005 |
| Cerebellum | 3853 | 0.672 | 0.0509 | 0.253 | 0.0192 | 0.005 |
| Cortex | 3852 | 0.6726 | 0.0509 | 0.2524 | 0.0191 | 0.005 |
| Frontal cortex (BA9) | 3842 | 0.6672 | 0.0501 | 0.2583 | 0.0194 | 0.005 |
| Hippocampus | 3847 | 0.6733 | 0.0507 | 0.252 | 0.019 | 0.005 |
| Hypothalamus | 3842 | 0.6615 | 0.0497 | 0.264 | 0.0198 | 0.005 |
| Putamen (basal ganglia) | 3852 | 0.6651 | 0.0503 | 0.2598 | 0.0197 | 0.005 |
| Amygdala | 3829 | 0.6679 | 0.0492 | 0.2589 | 0.0191 | 0.0049 |
| Spinal cord - cervical C-1 | 3771 | 0.6739 | 0.0475 | 0.2558 | 0.018 | 0.0048 |
| Substantia nigra | 3834 | 0.6736 | 0.0482 | 0.2552 | 0.0182 | 0.0048 |

Abbreviations: SNP, single nucleotide polymorphism

## 4. Discussion

In this analysis, we found neither common nor rare variants within *SLC7A11* to be associated with PD across the AMP-PD and UK Biobank cohorts, and similarly, observed no associations with GCase enzymatic activity. We identified 11 variants near or within the SLC7A11 locus that were significantly associated with gene expression across multiple brain tissues, yet these eQTL signals did not overlap with any PD GWAS signals in our colocalization analyses, suggesting that neither genetic variation nor gene expression of SLC7A11 have an important role in PD.

A relationship between *SLC7A11* and PD has been suggested in recent studies, demonstrating that dysfunction in this gene may influence PD pathogenesis at the cellular level. *SLC7A11* deficiency can result in lysosomal dysfunction and oxidative stress, facilitating alpha-synuclein accumulation, the hallmark pathology of PD, as demonstrated in SLC7A11-knockout mouse primary neurons and human iPSC-derived dopaminergic neurons from PD patients ^5^. Similarly, it was suggested that SLC7A11 may affect PD through ferroptosis, with hypoxia-inducible factor-1 (HIF1-) positively regulating SLC7A11 to inhibit this process ^13^. It was further demonstrated that upregulation of SLC7A11 can suppress microglial ferroptosis and reduce alpha-synuclein accumulation ^14^. In addition, epigenetic studies have identified DNA methylation changes at the SLC7A11 locus to be associated with PD, suggesting a potential regulatory role for this gene in disease-related pathways ^15^. Since our genetic and expression results do not support the previous functional results described above, further studies are needed to determine whether *SLC7A11* is involved in PD. Specifically, since variants that increase or reduce the expression of SLC7A11 are not associated with PD across all the brain regions we analyzed, it might imply that up-or down-regulation SLC7A11 have no meaningful role in PD.

This study has several limitations. First, differences in cohort design, sampling strategies, sequencing platforms, and variant-calling pipelines across datasets (AMP-PD and UK Biobank) may have introduced variability in variant detection and association estimates. These differences, including recruitment strategies, diagnostic criteria, and quality-control thresholds, may influence the number and type of variants identified and potentially obscure subtle genetic effects or introduce bias. However, our meta-analysis approach mitigates some of these issues.

Second, the rare variant burden analyses and the common variant analyses were restricted to individuals of European ancestry to minimize population stratification, which may limit the generalizability of these findings to other populations. Further studies including more diverse populations are needed to validate these results. Third, although our study leverages large publicly available datasets, it is limited by the use of heterogeneous sequencing approaches and reliance on summary-level data, which precludes detailed functional assessment. Finally, while our results identify regulatory associations within the SLC7A11 locus, they do not establish functional consequences, and additional studies will be required to determine how these variants affect gene expression or cellular function.

## 5. Conclusion

In conclusion, our findings do not support *SLC7A11* as a genetic susceptibility locus for PD. Although functional studies demonstrate that perturbation of SLC7A11 influences cellular pathways implicated in PD pathogenesis. Variations in this gene, including variants with strong effects on gene expression in different brain regions, does not appear to meaningfully alter PD risk. However, our analyses specifically address disease susceptibility and do not evaluate potential effects on progression or clinical heterogeneity. Further studies integrating genetic and longitudinal clinical data are needed to determine whether variation at this locus influences PD progression or phenotypic variability rather than disease susceptibility, and whether non-genetic mechanisms such as epigenetic regulation contribute to its role in PD.

## Acknowledgements

We would like to express our sincere gratitude to all participants, investigators, and research teams whose contributions made this study possible. We thank the UK Biobank and Accelerating Medicines Partnership Parkinson’s Disease (AMP-PD) cohorts for providing the data used in this work. This study used data from the UK Biobank under Application Number 45551 and data was obtained from the AMP-PD Knowledge platform. AMP-PD is a public-private partnership managed by the Foundation for the National Institutes of Health and supported by the National Institute of Neurological Disorders and Stroke, the Aligning Science Across Parkinson’s Initiative, The Michael J. Fox Foundation for Parkinson’s Research, and industry partners. We also acknowledge the Global Parkinson’s Genetics Program (GP2), funded by the Aligning Science Across Parkinson’s initiative and implemented by The Michael J Fox Foundation for Parkinson’s Research, for providing the data used in this study.

Computational analyses performed in this study were supported by NeuroHub, Calcul Québec, and the Digital Research Alliance of Canada, with additional support from the Canada First Research Excellence Fund through the Healthy Brains, Healthy Lives initiative at McGill University.

We would additionally like to acknowledge the support from G-Can, the GBA1-Canada initiative, an open-science platform aimed at advancing GBA1 research, with support from the Hilary and Galen Weston Foundation, the Silverstein Foundation, and J. Sebastian van Berkom and Ghislaine Saucier.

## STATEMENTS & DECLARATIONS

### Ethical Consideration

Not applicable

### Consent to Participate

Not applicable.

### Consent for Publication

Not applicable.

### Author Contributions

Z. G.-O. conceived the main idea of this study. Y.L. performed the data acquisition, analyses, investigations, as well as the preparation and visualisation of the initial draft. S.C.P., E.S. and K.S. contributed to the methodology, data acquisition and data analysis. Z.G.-O. also provided critical revisions and supervision of the project. All authors were involved in the revision and approval of the final version of the manuscript.

### Data Availability

For the analysis of rare variants, both AMP-PD Knowledge Platform and the UKBB datasets require approval by the corresponding institution for access. For the common variant analysis, the summary GWAS statistics can be accessed by the public. All data significant to this study can be found in the Tables section or in the Supplementary Data section of this paper.

